# Wellbeing After Stroke (WAterS): feasibility testing of a co-developed Acceptance and Commitment Therapy (ACT) intervention, to support psychological adjustment after stroke

**DOI:** 10.1101/2023.10.13.23296276

**Authors:** Emma Patchwood, Hannah Foote, Andy Vail, Sarah Cotterill, Geoff Hill, members of the WAterS PCPI Group, Audrey Bowen

## Abstract

**Objective:** Feasibility test a co-developed intervention based on Acceptance and Commitment Therapy (ACT) to support psychological adjustment post-stroke, delivered by a workforce with community in-reach.

**Design:** Observational feasibility study utilising Patient, Carer, Public Involvement (PCPI).

**Setting:** Online. UK.

**Participants:** Stroke survivors with self-reported psychological distress 4+ months post-stroke

**Interventions:** The co-developed Wellbeing After Stroke (WAterS) intervention includes: nine weekly, structured, online, group sessions for stroke survivors, delivered via a training programme to upskill staff without previous ACT experience, under Clinical Psychology supervision.

**Main measures:** Feasibility of recruitment and retention; data quality from candidate measures; safety. Clinical and demographic information at baseline; Patient Reported Outcome Measures (PROMs) via online surveys (baseline, pre- and post-intervention, 3 and 6 months after intervention end) including Mood (HADS), Wellbeing (ONS4), Health-Related Quality of Life (EQ5D5L), Psychological Flexibility (AAQ-ABI) and Values-Based Living (VQ).

**Results:** We trained eight staff and recruited 17 stroke survivors with mild-to-moderate cognitive and communication difficulties. 12/17 (71%) joined three intervention groups with 98% attendance and no related adverse events. PROMS data were well-completed. The HADS is a possible future primary outcome (self-reported depression lower on average by 1.3 points: 8.5 pre-group to 7.1 at 3 month follow up; 95% CI 0.4 to 3.2).

**Conclusion:** The WAterS intervention warrants further research evaluation. Staff can be trained and upskilled to deliver. It appears safe and feasible to deliver online to groups, and study recruitment and data collection are feasible. Funding has been secured to further develop the intervention, considering implementation and health equality.

## Introduction

Up to 75% of stroke survivors report changes to their mental health including psychological distress, depression, and anxiety, for which they report inadequate support [1, 2]. Supporting psychological adjustment is the number one priority for Life After Stroke research in the UK [3–5] but we lack evidence-based interventions and there is a severely under-resourced applied psychological clinician workforce in the UK (e.g., Clinical Psychologists)[6]. However, psychological interventions to support life after stroke could also be provided by other professionals, which may prevent the development of more severe difficulties with mood in Life After Stroke [7].

Acceptance and Commitment Therapy (ACT) is a third-wave, transdiagnostic, cognitive-behavioural psychotherapy [8]. It focuses on supporting clients to develop a more psychologically flexible relationship with the present moment and any associated challenges, towards committed action in service of personal values. ACT has potential benefits for supporting psychological wellbeing and preventing depression after stroke and acquired brain injury [9–17]. It has been delivered as a group-based face-to-face intervention for stroke survivors [18]; group delivery may appeal to under-resourced healthcare systems [19, 20]. In addition, remotely delivered interventions, rather than face-face, may be an optimised method of service delivery when clinically indicated. The COVID-19 pandemic caused substantial disruption to normal health services, with telemedicine broadly recommended as a result [21]. Guidance within the UK National Health Service (NHS) suggests that remote consultations may be suitable for stroke support groups, group rehabilitation and counselling (or similar) services [22] There is a strong case for research into the acceptability and feasibility of remote service delivery models.

In close collaboration with a study-specific Patient Carer and Public Involvement (PCPI) advisory research group and other experts, we co-developed the Wellbeing After Stroke (WATerS) intervention between January 2020 and May 2021, taking longer than anticipated due to the pandemic. WAterS built on an existing NHS intervention called Living Well with Neurological Conditions [23] and is a structured ACT-informed intervention delivered online to groups of stroke survivors using Zoom. An adjunct staff training and support programme was also developed to upskill staff without previous experience in this therapy with the skills and knowledge to safely deliver WAterS (for detailed intervention description see Methods and Appendix 1).

This paper reports results of the Wellbeing After Stroke (WAterS) feasibility study which had the following specific objectives:

1. Evaluate the feasibility of delivering the WAterS intervention: both the staff training and the stroke survivor intervention;
2. Explore the feasibility of a future research trial, including: recruitment and retention of stroke survivors; inclusion/exclusion criteria; data quality from baseline and candidate outcome measures; adverse events;

A process evaluation, exploring fidelity and acceptability utilising qualitative and quantitative data, will be reported separately.

## Methods

This study included methodological components from the development and feasibility phases of the Medical Research Council’s new framework for developing and evaluating complex interventions [24], and collaborative-level patient involvement [25]. Ethics approval was secured from the University of Manchester (ref: 2021-11134-18220) who also acted as sponsor. The study is registered: https://clinicaltrials.gov/ct2/show/NCT04655937

A study-specific PCPI group of four individuals with experience of stroke and caring was set up at the early stages of the study. They met regularly to support initial intervention development, including providing feedback on proposed intervention materials and role-playing the intervention in practice. They advised on participant recruitment and data collection, considering burden for stroke survivors.

The research team members responsible for all participant-facing procedures were EP (10 years’ experience in applied stroke research) and HF (PhD candidate and Speech and Language Therapist). Due to the remote delivery, no geographical limits on recruitment in the UK were in place; all research procedures were conducted by telephone or Zoom.

Staff participants were recruited to facilitate delivery of the intervention, with two staff per intervention session deemed necessary to ensure patient safety, cover unexpected absence and ensure smooth running of the technology. They worked for a major UK charity specialising in stroke (Stroke Association) and were eligible if they had: at least 6 months service working with stroke survivors; capacity and time to dedicate to the study; experience facilitating groups; no prior experience using ACT as a therapeutic model. Staff were opportunistically sampled (identified by their senior leaders) and those eligible were provided with study information prior to informed consent, with training commencing immediately after recruitment.

The initial staff training programme (see Appendix 1) was delivered online using Zoom over four weekly half-day sessions, led by a Clinical Psychologist. After training, support for staff consisted of an online weekly one-hour group session with a Clinical Psychologist (GH) providing opportunities to reflect on the therapy and troubleshoot the protocol with an expert and each other.

Following completion of staff training, stroke survivor participants were recruited for feasibility testing. The eligibility criteria were adults (aged 18+) at least 4 months post-stroke, with self-reported difficulties adjusting to their stroke, internet access and sufficient English language (and willingness) to engage in a remotely-delivered intervention. Study advertisements were circulated via third sector, research networks and social media, with interested parties self-referring to the research team for information and consent. Eligibility was determined based on self-report and research team clinical judgement. The intervention was stroke survivor focused, but informal carers (e.g. family members or friends) could be invited to support, on request of the stroke survivors. No formal data were collected about informal carers but a procedure for involving informal carers was in place in the interests of data protection, regulation and safety.

Baseline data on staff participants included demographic and employment related data. Baseline data on stroke survivors included demographic and clinical data as well as Patient Reported Outcome Measures (PROMs). During a Zoom call at baseline, the latter were asked to self-rate: communication; reading; writing; memory; processing; and mental health (from 0 = very poor to 10 = very good). Then researcher-administered assessments were completed: components of the Montreal Cognitive Assessment (MoCA)[26], Frenchay Aphasia Screening Test (FAST)[27] and the Quick Aphasia Battery for Comprehension (QAB-C)[28]. Following this Zoom session, researchers rated stroke survivor participants on the Therapy Outcome Measures (TOMS)[29] scales for aphasia (activity level) and cognition (impairment level). Participants were then sent a link to an online survey platform to self-complete baseline measures related to: mood (the Hospital Anxiety and Depression Scale (HADS))[30]; personal wellbeing (ONS4) [31]; independence in activities of daily living (the Modified Barthel Index (mBI)) [32]. We also asked questions about other support they were receiving for their mental health and wellbeing. Participants had the option of telephone support from a researcher to complete online measures.

We had capacity to run up to four intervention groups. This was not a randomised controlled trial and there was no control group. As a feasibility study, we did not have a pre-determined sample size but anticipated a minimum of four stroke survivors per group (maximum of eight). After sufficient participants were recruited to run groups, stroke survivors had the choice of joining or declining the intervention. If they declined, they could either leave the study completely or continue completing online surveys at regular time intervals (see Table 1), to explore the feasibility of data collection in the absence of intervention. Once sufficient numbers agreed to intervention they were randomly assigned to therapy groups by researcher EP. Participant IDs were sorted according to a random number generator with equal number of participants assigned to the intervention groups (labelled Group A, B, and so on).

**Table 1:** summary of stroke survivor data collection timepoints.

| **Data / measures collected** | **Baseline** | **Pre-group** | **Post-group** | **3 month follow up** | **6 month follow up** |
| --- | --- | --- | --- | --- | --- |
| **Demographic and clinical data (see text)** | X |  |  |  |  |
| **Mood (HADS)** | X | X | X | X | X |
| **Wellbeing (ONS4)** | X | X | X | X | X |
| **Health-related quality of life (EQ-5D-5L)** |  | X | X | X | X |
| **Psychological flexibility (AAQ-ABI)** |  | X | X | X | X |
| **Values-based living (VQ)** |  | X | X | X | X |
| **Questions on additional mental health support being received** | X | X | X | X | X |

The intervention involved ACT-informed activities delivered over nine weekly sessions online using Zoom. A client handbook was sent to participants in advance of intervention commencement to support engagement. The handbook included written summaries of the session content, worksheets and homework to complete between weekly sessions. Each session was facilitated by two trained staff: one designated ‘lead’ to have primary responsibility for delivering content, with the other designated ‘support’ to ensure smooth flowing of the session, including technical and safety issues. Support staff could lead on components of sessions, depending on confidence and the lead/support dynamic. The intervention was highly structured (see Appendix 1) to enhance fidelity and equip non-experts to facilitate.

Table 1 summarises the additional data collected through online surveys from stroke survivor participants at each timepoint: immediately prior to intervention group (pre-group); after final intervention group session (post-group); and 3- and 6-months later (follow ups). These data included repetition of some baseline measures as well as others exploring health-related quality of life (EQ-5D-5L) [33], and two questionnaires to explore the key targeted processes of change in this psychotherapy: psychological flexibility (AAQ-ABI)[34]; and values-based living (VQ)[35]. They also included a question to explore any additional mental health support being accessed at that time.

This study was not powered to examine effectiveness. Outcomes were collected to explore feasibility of data collection and inform the choice of a primary outcome for an eventual trial. Serious Adverse Events (SAEs) were collected as they occurred. Figure 1 shows a summary of the study processes and timelines.

**Figure 1.**
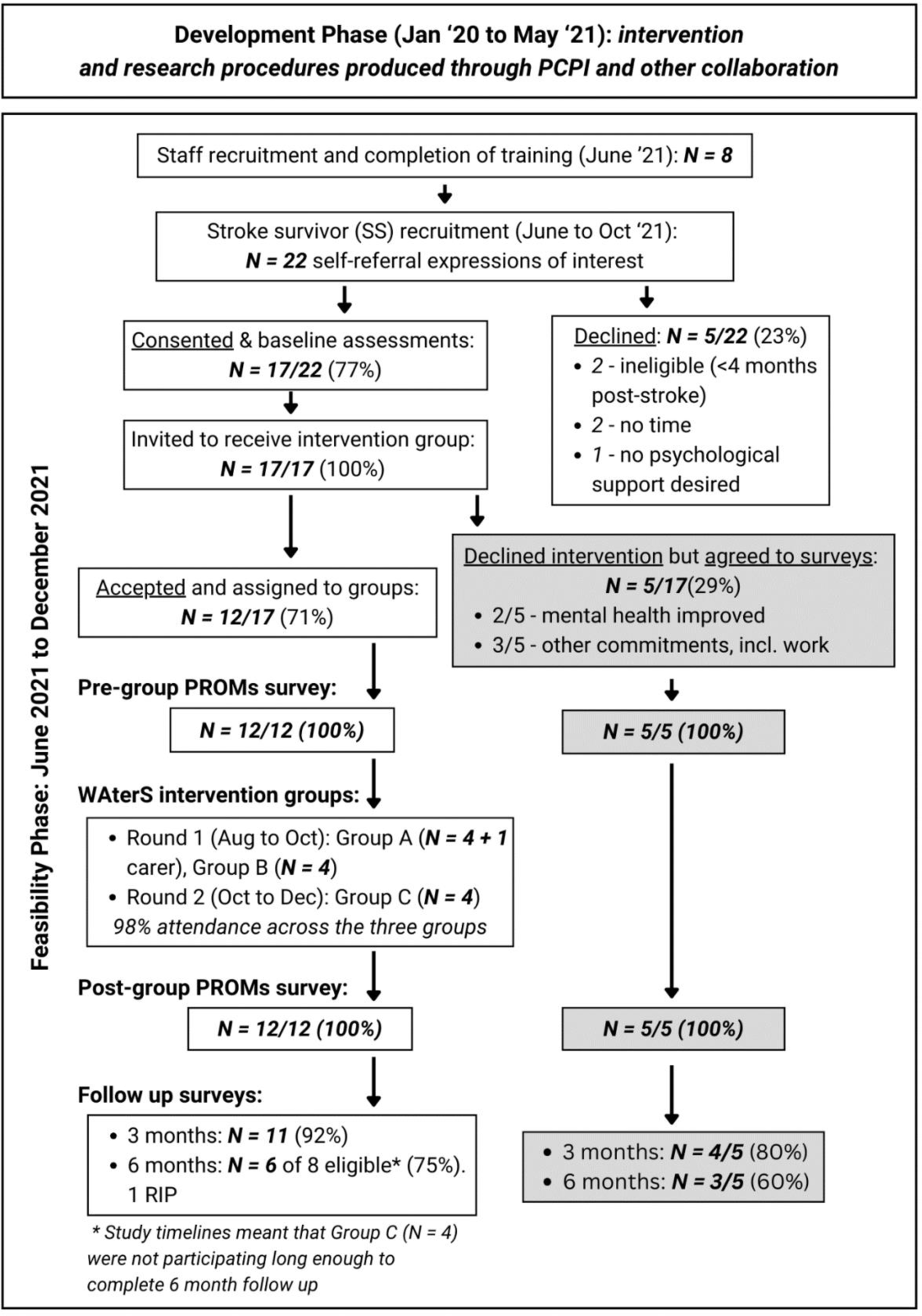
Study timelines and flow of participants.

## Results

Recruitment and testing began formally in June 2021. Figure 1 shows the flow of participants through this phase of the study, as well as study timelines.

It proved feasible to recruit and train eight staff. All were female, with a mean age of 53 (SD: 7.63) and mean number of years working for the organisation of 6 (range 1-15). Five staff were trained as lead practitioners, and three as support. All practitioners had the required experience and, in addition, five had a UK level four counselling qualification and were designated as the ‘lead’ facilitators. Seven of the eight then supported delivery of the intervention.

Recruitment of stroke survivors appeared feasible using our range of methods, amidst an ongoing pandemic. From 22 expressions of interest, 17 (77%) consented and completed baseline assessments, with one informal carer also involved to support their spouse. When invited to receive intervention therapy groups, 12/17 (71%) accepted and 5/17 (29%) declined groups but agreed to continue completing surveys. Table 2 shows stroke survivor participant characteristics at baseline; separate columns show data for 12 who participated in intervention vs the 5 survey-only.

**Table 2.**
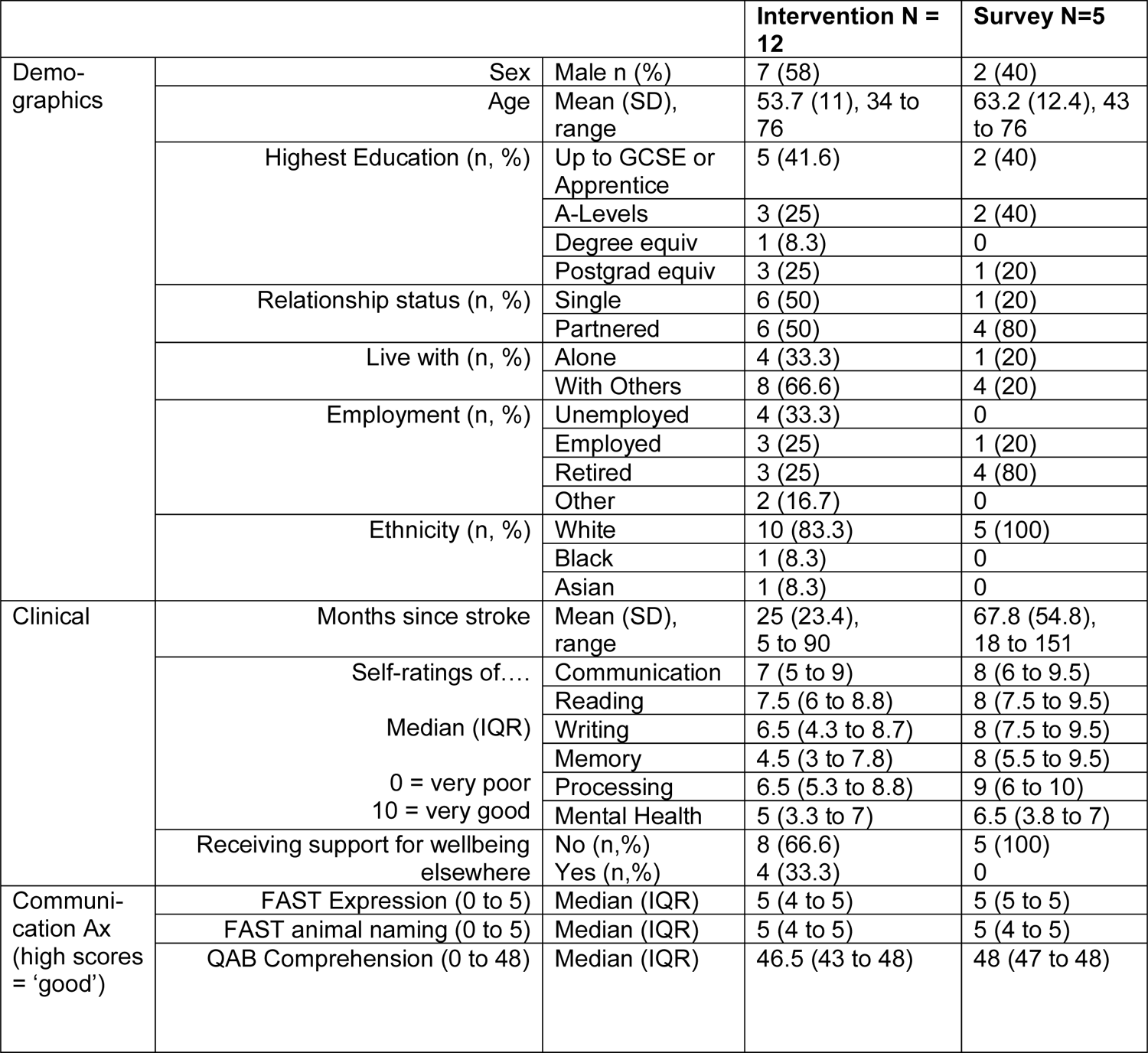
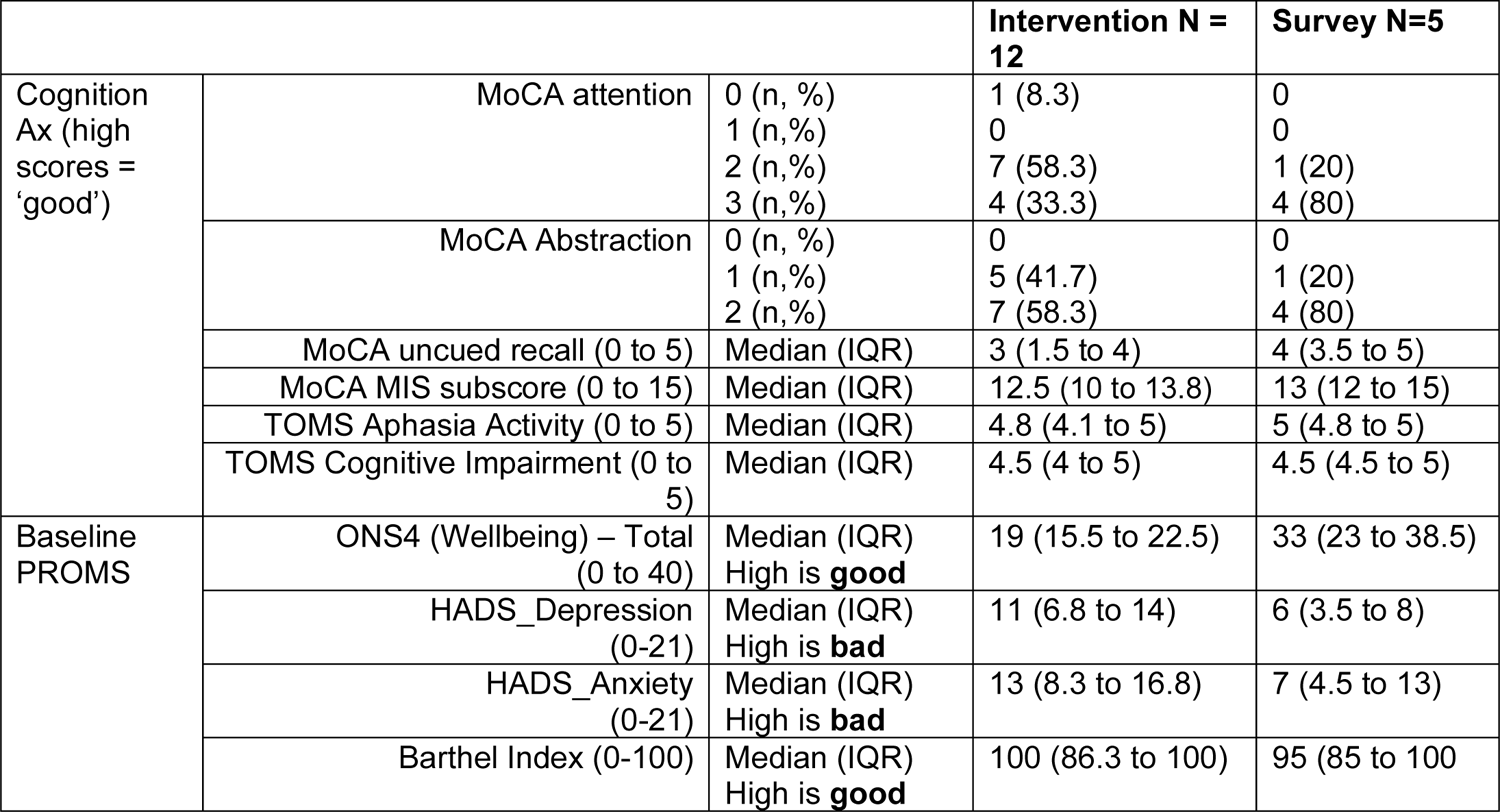
Baseline characteristics: intervention participants (N=12); and survey only (N=5)

Roughly half the stroke survivor participants were male and the majority were White British. Of the 12 who joined the intervention, the mean (SD) age was 53.7 (11) years and mean (SD) months post-stroke was 25 (23.4). Other measured characteristics indicate that all participants had mild-moderate cognitive and communication difficulties (only 2/12 intervention participants self-reported expressive aphasia).

The 12 participants consenting to intervention were assigned to three WAterS groups: A, B, C. The 5 who declined intervention appeared to be later post-stroke, were all receiving other support and had less self-reported need. The mean (SD) wait time from consent to intervention start was 31 (14.5) days (min = 14; max = 68). All group sessions happened weekly over 9 weeks as planned, with excellent attendance at 98%. No serious adverse events related to the intervention were recorded. The experiential exercises did evoke emotional responses from participants, including distress, but these were managed by the group facilitators.

Table 3 shows data for intervention group (N=12) and survey-only cohort (N=5) collected at the four timepoints after baseline (pre- and post-group; 3- and 6-month follow up), including numbers to show attrition. Online data collection from stroke survivors appeared feasible for a future trial. The mean (SD) time to complete surveys was 19.3 (19.1) minutes (min = 4.1; max = 90). The 90 minute maximum was when a researcher supported completion (only required for one participant). Fully completed datasets with no missing items were captured at pre- and post-group timepoints. At 3 month follow up, completed datasets were 80-92% across the sample; 6-month follow up recorded 60-86% completed datasets.

**Table 3.** PROMs data for 4x additional timepoints. Intervention (N=12); and surveys only (N=5)

|  |  | Intervention (N = 12) + |  | Surveys-only (N=5) ++ |  |
| --- | --- | --- | --- | --- | --- |
|  |  | Mean (SD) | Change from Pre-group (95% CI) <sup>□</sup> | Mean (SD) | Change from Pre-group (95% CI) <sup>□</sup> |
| ONS4<br>(0 to 40; High is good) | Pre | 20.4 (7.9) |  | 24 (3.9) |  |
|  | Post | 20.3 (6.3) | -0.2 (-4.5 to 4.8) | 25 (8.2) | -1 (-12.4 to 10.4) |
|  | 3 month | 20.0 (4.4) | -1.1 (-4.4 to 2.2) | 31.2 (6.7) | -7.3 (-17.9 to 3.4) |
|  | 6 month* | 19.0 (6.8) | -2.3 (-10.7 to 6.1) | 33 (7.8) | -9.3 -27 to 8.3) |
| HADS (Depression)<br>(0-21; High is bad) | Pre | 8.4 (4.7) |  | 7.2 (2.8) |  |
|  | Post | 8.3 (5.3) | 0.2 (-1 to 1.4) | 8.6 (5.7) | -1.4 (-7.0 to 4.2) |
|  | 3 month | 7.1 (4.5) | 1.4 (-0.4 to 3.2) | 5.8 (2) | 1.3 (-3.3 to 5.8) |
|  | 6 month* | 9.8 (5.9) | 1.5 (-1.7 to 4.7) | 5 (1.4) | 1.5 (-30.3 to 33.3) |
| HADS (Anxiety)<br>(0-21; High is bad) | Pre | 11.3 (3.6) |  | 7.8 (3.5) |  |
|  | Post | 10.5 (4.6) | 0.8 (-0.9 to 2.4) | 10.4 (5.7) | -2.6 (-8.5 to 3.3) |
|  | 3 month | 11.3 (5.5) | 0.2 (-2.3 to 2.7) | 8 (5.4) | 0.3 (-3.5 to 4.0) |
|  | 6 month* | 11.5 (2.6) | 2 (-1.7 to 5.7) | 4.33 (3.2) | 2.3 (-3.9 to 8.6) |
| EQ-5D-5L<br>Health Utility<br>(-1 to +1; High is good) | Pre | 0.57 (.28) |  | 0.63 (.17) |  |
|  | Post | 0.53 (.32) | 0.03 (-0.1 to 0.1) | 0.65 (.11) | -0.02 (-0.1 to 0.1) |
|  | 3 month | 0.50 (.26) | -0.0 (-0.1 to 0.1) | 0.58 (.19) | 0.04 (-0.1 to 0.1) |
|  | 6 month* | 0.58 (.26) | -0.1 (-0.2 to -0.0) | 0.54 (.15) | 0.02 (-0.1 to 0.2) |
| AAQ-ABI<br>(0 to 36; high is good /<br>greater flexibility) | Pre | 15.9 (5.8) |  | 14.4 (6.9) |  |
|  | Post | 16.3 (7.9) | -0.4 (-3.9 to 3.2) | 14.6 (8.3) | -0.2 (-5.9 to 5.5) |
|  | 3 month | 15.3 (8.1) | 0.5 (-3.9 to 5.0) | 14 (6.3) | -1.3 (-9.1 to 6.6) |
|  | 6 month* | 12.3 (8.9) | 4.3 (-4.8 to 13.5) | 8.3 (8.5) | 3 (0.52 to 5.5) |
| VQ (Progress)<br>(0-30; High is good /<br>alignment with values) | Pre | 18.3 (5.8) |  | 21.2 (7.5) |  |
|  | Post | 16.7 (8.4) | 1.6 (-0.8 to 3.9) | 17.8 (6.2) | 3.4 (-4.7 to 11.5) |
|  | 3 month | 16.4 (7.2) | 0.7 (-3.0 to 4.4) | 22.3 (7.2) | 1.3 (-3.5 to 6.0) |
|  | 6 month* | 14.7 (8.2) | 1.5 (-6.1 to 9.1) | 25.2 (8.1) | -1 (-5.3 to 3.3) |
| VQ (Obstruct)<br>(0-30; High is bad /<br>interference with values) | Pre | 16.8 (6.2) |  | 10.4 (6.7) |  |
|  | Post | 15.5 (6.6) | 1.3 (-2.2 to 4.8) | 16.8 (4.1) | -6.4 (-13.7 to 0.9) |
|  | 3 month | 17.7 (6) | -0.4 (-3.5 to 2.7) | 11 (11.6) | -1.0 (-8.5 to 6.5) |
|  | 6 month* | 17.33 (8.9) | 0.2 (-6.9 to 7.2) | 7.3 (8.1) | 0.0 (-4.3 to 4.3) |
| Receiving support for<br>wellbeing elsewhere<br>No or Yes: n (%) | Pre | No: 3 (25) | Yes: 9 (75) | No: 4 (80) | Yes: 1 (20) |
|  | Post | No: 6 (50) | Yes: 6 (50) | No: 4 (80) | Yes: 1 (20) |
|  | 3 month | No: 5 (50) | Yes: 5 (50) | No: 3 (75) | Yes: 1 (25) |
|  | 6 month* | No: 3 (50) | Yes: 3 (50) | No: 2 (67) | Yes: 1 (23) |

+ all N= 12 completed pre- and post- group, n=9 to 11 completed at 3-months, n=6 at 6-months
++ all N=5 completed pre- and post- group, n=4 completed at 3-months, n=3 at 6-months
<sup>□</sup> Numbers at each follow-up point vary. Change scores are for those with valid data for each outcome.
\* Study timelines meant that Group C (N=4) were not participating long enough to complete 6 month follow up. 1 participant died between 3- and 6-month follow up.

## Discussion

This study demonstrated the feasibility of delivering the WAterS online, well-being group intervention, and of its evaluation using online data collection. Accessibility to the intervention, and the study, benefitted from a dedicated, study-specific PCPI panel. The intervention was feasible to deliver using a third sector workforce with no previous experience of ACT, following the WAterS staff training programme and supervision from an NHS Clinical Psychologist. Study recruitment was successful, despite the pandemic, but could be enhanced in future work. Attendance at the online groups (98%) was excellent for those who opted in, and future work will focus on widening access, mindful of health inequalities.

The routes to recruitment for stroke survivors required self-referral following advertisement. We have useful descriptive data on those who accepted intervention, and those who declined intervention but agreed to provide follow up data. Typically, the latter reported less need which speaks to the validity of our data. We don’t have data on the proportion of people who saw the adverts, were potentially eligible, and did not self-refer. Understanding rates of eligibility in a population helps power and inform future trials [36], and the methods of recruitment used in this study may need to be enhanced. Utilising stroke healthcare pathways to recruit through healthcare professionals may be a useful added strategy for future work, whilst mindful of potential gatekeeping. However, study recruitment using social media and digital platforms, including for trials, is becoming more common [37]. It can increase reach to certain communities, including those likely to opt into an online intervention, but is threatened by digital poverty and may result in a sample that is skewed towards lower stroke morbidity and higher social economic status [38, 39]. We recruited a predominantly white (83%), well-educated, mild-to-moderate morbidity sample. This racial profile is similar to existing studies in stroke but reflects issues with equity of access, considering the well-documented health inequalities in stroke [40, 41]. Future work must carefully consider how to engage a more diverse range of stroke survivors. This would include those with aphasia, for whom English is not their first language, and those with different cultural expectations of therapy and of participating in research.

All 12 intervention group participants – including the two with self-reported (and observable) expressive aphasia - were able to engage in the intervention. Eligibility was explored via self-report and researcher judgement, without use of cut-off scores on assessments of mood or cognition. Our data simply describe who took part and are not sufficient to comment, post-hoc, on whom this intervention might be best-suited. Offering ACT to individuals with sub-threshold psychological disorders may help *prevent* mental health crises [13, 14]. Assessing perceived readiness or willingness to engage in this kind of psychotherapy may be more important than assessing mood or impairment when considering eligibility [42, 43]. The choice of researcher-administered baseline assessments were pragmatically chosen based on a number of factors including: explorations of acceptability and burden with the study PCPI group; ease of remote administration with no special equipment required; ease of access to obtain tools; researcher and clinical judgement. Fully remote, computerised assessments of cognition could be explored in future work for describing baseline profiles, although more research is required to validate these approaches in stroke [44, 45].

As anticipated for a mental health intervention, the experiential exercises evoked some emotional responses from participants. The facilitators provided emotional support (e.g. supported exploration of the triggering experiences) and the intervention appeared safe in this study. It is important not to conflate safety with ‘no distress experienced’; participants sometimes reflected that potentially unwanted emotional responses could lead to adjustment and positive transformation. Qualitative interview data exploring stroke survivor experiences in detail are discussed elsewhere (paper in progress). We had not anticipated recruiting staff with counselling qualifications to facilitate (it was not an original staff eligibility criterion) but the added counselling skills of lead staff may have contributed to the safety in this trial. Exploring the minimum requirements for staff to deliver an intervention like WAterS safely - and with fidelity - is an avenue for future research.

Five stroke survivor participants (29%) declined the intervention but completed online outcomes surveys; two due to self-reported mental health improvements and three due to lack of availability to attend the therapy sessions (see Fig 1). The latter barrier may have been overcome with pre-determined dates and times for intervention discussed during recruitment. The former is interesting, since all recruited participants initially reported difficulties with their mental health and a desire to receive support. However, their baseline data show lower self-reported anxiety and depression levels in these five compared to the 12 who opted to join a group. Future work could explore the value of providing a little more time for stroke survivors to decide whether their self-reported needs warrant intervention. Space to normalise their feelings may be sufficient for some, and preferential to the treatment burden of a nine-week intervention that others with greater needs deem appropriate to take on. The delivery and evaluation of any intervention following stroke must acknowledge the number of impairments that can result, and that many stroke survivors will also be managing multiple long term conditions [46].

Online data collection appeared acceptable and feasible in this trial. Our choice of outcomes was informed by the literature [47] and PCPI. This small study was not powered to explore measurement outcomes. All change scores in the data must be viewed with caution, and the tiny numbers at six months mean we have no useful data at that timepoint. However, HADS Depression may be a candidate measure for primary outcome in future research; it was lower on average by 1.4 points from pre-group to 3 month follow up (8.5 to 7.1; 95% CI −0.4 to 3.2). Minimal clinically important difference (MCID) on HADS are not available for stroke to our knowledge, but research in cardiovascular disease suggests MCID of 1.7 points [48]. In addition, other studies using this therapeutic model in acquired brain injury have suggested that the HADS is sensitive to change [10, 11].

Psychological flexibility and values-based living were relatively unchanged during the three month follow up period with wide confidence intervals in our small sample size. However, as key hypothesised psychological constructs targeted by ACT, they may be important to continue to include in future research in order to derive and test a theory of change related to if/how this intervention influences outcomes [49].

In conclusion, the co-developed online WAterS group intervention was feasible to deliver to stroke survivors using a trained workforce, and had a high adherence rate. Further in-depth data on fidelity and acceptability will be reported in a separate paper. Success with study processes, online data collection etc, suggested further research evaluation is feasible and warranted with enhancements to recruitment routes and outcome measures. Further funding has been secured to develop the clinical and research protocols for WAterS, focused on addressing aspects of health inequalities and widening the workforce. This future funding will gather more data to inform the choice of baseline and outcome measures and ultimately work towards a robust randomised controlled trial with process evaluation.

### Clinical Messages

- The ACT-informed WAterS intervention is feasible to deliver remotely to groups of stroke survivors in the context of a research study.
- Staff without ACT expertise can feasibly be recruited and trained to deliver WAterS.
- HADS depression may be a candidate primary outcome for trials exploring effectiveness of WAterS.

## Data Availability

All data produced in the present study are available upon reasonable request to the authors.

## Acknowledgements

This independent research was funded by a Stroke Association Postdoctoral Fellowship Award (Ref SA PDF 18100024). The views expressed are those of the author(s) and not necessarily those of the Stroke Association. Authors also acknowledge funding from The University of Manchester Research Impact Scholarship (MC). Funders had no role in study design, execution, analysis or results interpretation. The authors extend thanks to patient participants, therapy and research staff, and our dedicated PCPI patient advisory group. Our PCPI group members are listed as co-authors and include: Mrs Ann Bamford, Mr Stephen Taylor, Mr Nigel Bamford, Ms Loretta Hanley. The authors declare no conflicts of interest.

---

**Appendix 1.** Description of WAterS intervention, including staff training programmed Based on adapted Template for Intervention Description and Replication (TIDieR) [50, 51.

|  |  | **Modifications – changes made during intervention development** (what, why, when, and how). |
| --- | --- | --- |
| **1. Brief name**<br>Provide the name or a phrase that describes the intervention | **Wellbeing After Stroke (WAtErS)**<br>Acceptance and Commitment Therapy (ACT) delivered remotely to groups of community-based stroke survivors |  |
| **2. Why**<br>Describe any rationale, theory, or goal of the elements essential to the intervention | Acceptance and Commitment Therapy (ACT) is a third wave transdiagnostic cognitive-behavioural therapy[8] that supports clients to adjust to current experiences and 'commit' to behaviours that are congruent with their personal values, in order to promote psychological well-being and prevent future mental health crisis.<br>The principles of ACT are advocated for patients with acquired brain injury, including stroke,[12-14] but the evidence base is limited. ACT interventions, including mindfulness components, can improve quality of life and psychological well-being for these clients[52], including those with aphasia [53].<br>We aimed to develop an ACT-informed psychotherapeutic intervention to support post-stroke adjustment and prevent mental health crises. The content was developed for stroke and resultant cognitive and communication difficulties through the use of relevant examples, audio-visual resources and provision of a client handbook.<br>In addition, we wanted the intervention to be deliverable by an available workforce to ensure reach if effective. As such, we also developed a staff training programme to equip professionals without previous ACT-expertise to deliver the intervention. | We began with an existing ACT intervention used in the NHS called Living Well with Neurological Conditions (LWNC) [17, 23] and – through working with the original developer of that course (author GH) and with PCPI input – we co-developed a more structured programme; tailored for stroke and script-informed to support delivery by non-experts<br>This 'modifications' column is primarily used to detail how co-development (including PCPI) adapted the intervention, described in detail in central column. |
| **3. What – material**<br>Materials: Describe any physical or informational materials used in the intervention, including those provided to participants or used in intervention | WAtErS consisted of several material components that are briefly stated here with more details in section 4 below:<br>1) Clinical protocol: detailing the delivery of 9 structured ACT-informed weekly intervention sessions for stroke survivors. | These documents modified over time through co-development, including PCPI input and role-play, leading to |
| delivery or in training of intervention providers. Provide information on where the materials can be accessed (e.g. online appendix, URL). | 2) Staff training protocol: to train staff in delivering the Clinical Protocol;<br>3) Staff training handbook: sent to staff in advance of training sessions to support engagement and knowledge consolidation.<br>4) Participant handbook: sent to stroke survivors in advance of group intervention beginning, to improve engagement with weekly sessions and act as an aide memoire.<br>PowerPoint Slides: used in both the Staff Training and Clinical protocols.<br>Additional audio-visual resources: used to deliver components of the Clinical and Staff training protocols; specifically YouTube videos for guided mindfulness activities.<br>All protocols, handbooks, slides and audio-visual resources are not yet available, since they are still being developed and evaluated. We will make all materials freely available under license once WAters has been successfully clinically evaluated. | adaptations in activities, including wording, format (old versions are saved)<br>For example:<br>- The number of sessions was increased during development (from 8 to 9) to allow more time for the group to 'get to know' one another and feel comfortable in this setting.<br>- Informal carers / family members were not originally planned to attend the group but PCPI suggested this was vital to be doable if stroke survivors wanted it (particularly for support with the technical aspects of using Zoom)<br>- Images used in the Participant Handbook to support understanding were modified through PCPI |
| **4. What – procedures**<br>Procedures: Describe each of the procedures, activities, and/or processes used in the intervention, including any enabling or support activities. | Staff were trained prior to delivering the Clinical Protocol to Stroke Survivor research participants.<br>Staff Training protocol was delivered over 4 weekly half-day sessions online using Zoom, led by a Clinical Psychologist with ACT expertise and utilising audio-visual resources. Staff Training protocol includes the theoretical basis of ACT; experiential ACT exercises; role-play of facilitation; familiarisation with materials for each of the highly structured sessions in the Clinical Protocol used with stroke survivors; and homework suggestions between weekly sessions to support knowledge consolidation.<br>Staff training handbook was sent in advance of training sessions summarising session content. Additional crib sheets were included on the technical aspects of running groups with stroke survivors using video conferencing software, as well as distress protocols for keeping clients safe.<br>After training, staff were supported when delivering the Clinical Protocol to stroke survivors (see below). Support consisted of an online weekly one-hour group session |  |
|  | with the Clinical Psychologist who had trained them (GH); providing opportunities to reflect on ACT and troubleshoot the Clinical protocol with an expert and each other.<br>Clinical Protocol comprises detailed session plans for 9 highly-structured intervention sessions, led by two trained staff members (see 5 below). Stroke survivors can invite a carer/family/friend for support during sessions. The session content was adapted for stroke and resultant cognitive and communication difficulties, through the use of relevant examples, audio-visual resources and provision of a client handbook (described below). The Clinical Protocol was ACT-informed with sessions incorporating a variety of ACT experiential exercises including: mindfulness practices; activities to identify personal values; exploration of metaphors and other cognitive defusion techniques; activities to cultivate commitment to value-congruent goals. The intervention was also broadly grounded in neurorehabilitation principles with integrated psychoeducation on a range of post-stroke difficulties (e.g., memory, word-finding problems, fatigue) delivered in ACT-informed exercises. Flexible scripts are included for staff to utilise: designed to promote congruence to ACT and increase replicability / fidelity. There were opportunities for 'free-form' discussion throughout the sessions but these had guides, tips, prompts for staff to use to enhance fidelity to protocol and congruence to ACT.<br>The Client handbook – sent to stroke survivor participants in advance of intervention sessions starting - summarised session content and included worksheets for session activities and homework that could be completed in between weekly sessions to embed learning and knowledge. The handbook is designed to be kept as a memory aid after the intervention finishes. |  |
| **5. Who provided**<br>For each category of intervention provider (e.g. psychologist, nursing assistant), describe their expertise, background and any specific training given. | Provision of Staff Training and Support: As above, staff training and support was delivered by a Clinical Psychologist with substantial experience and expertise in ACT.<br>Provision of the Clinical Protocol to stroke survivors: Each of the nine weekly sessions were facilitated by two trained staff (with additional staff trained as 'backups' to cover absences). Of the two active staff running a therapy session, one would be designated 'lead' to have primary responsibility for delivering content, with a 'support' designated staff to ensure smooth flowing of the session, including technical issues. Support staff could lead on components of sessions, depending on | Changes within the Stroke Association (the organisation employing trained staff meant that Emotional Support staff (with Level 4 counselling qualification) were involved in the study and led in delivery. We had intended to have only 'lower grade' coordinators involved to explore whether |
|  | confidence and the lead/support dynamic. All trained staff were employed by the Stroke Association and had no previous experience of ACT. They were coordinators, experienced in working with people who have had a stroke and in facilitating groups. The 'leads' had this experience, and also a UK level four counselling qualification. | they could be upskilled to deliver this intervention. It is likely that the additional skills of the Emotional Support leads contributed to the success of the study. |
| **6. How**<br>Describe the modes of delivery (e.g. face-to-face or by some other mechanism, such as internet or telephone) of the intervention and whether it was provided individually or in a group. | Staff training was delivered by one Clinical Psychologist (as above) to a group of eight staff<br>The Clinical intervention was delivered across three groups in total:<br>- Two staff (one lead and one support) facilitated each group (as above) per group<br>- Four stroke survivors were included in each group, which one group including an one informal carer (spouse)<br>All sessions were delivered online using Zoom video-conferencing. | We had intended to run a face to face intervention but the move online was catalysed by the COVID-19 pandemic.<br>Group size dictated by recruitment but we had anticipated up to 8 stroke survivors per group. |
| **7. Where**<br>Describe the type(s) of location(s) where the intervention occurred, including any necessary infrastructure or relevant features. | This was an online course, so all trainers and facilitators were in their own homes/workplaces as appropriate for them. |  |
| **8. When and how much**<br>Describe the number of times the intervention was delivered and over what period of time including the number of sessions, their schedule, and their duration, intensity or dose | Staff training: 4 x weekly sessions of 4 hours duration, with breaks during sessions and homework in between weekly sessions.<br>Clinical Protocol for stroke survivors: 9 x weekly sessions of 2 hours planned duration with a 20 minute break and homework in between weekly sessions | Sessions with stroke survivors with the Clinical Protocol were not always 2 hours long. Some sessions were shorter but sessions often ran over by 10-30 minutes. |
| **9. Tailoring**<br>If the intervention was planned to be personalised, titrated or adapted, then describe what, why, when, and how | The Clinical intervention was protocolised and script-informed although there was scope for minor amounts of tailoring to fit the staff style of delivery. Going 'off-script' was allowed in the interests of supporting those in the 'room'<br>There were aspects of the intervention that the deliverer had more control over – leading group discussions/reflections was a key part of the intervention. This was down to the facilitator's skill with the group. |  |
| **10. How well – planned**<br>If intervention adherence or fidelity was assessed, describe how and by whom, and if any strategies were used to maintain or improve fidelity, describe them. | Adherence was assessed using a bespoke WAtErS fidelity tool, self-completed by staff, with a sub-set completed by researchers to enable investigation of agreement.<br>Fidelity to the process/mechanism of the intervention was assessed using the an ACT-specific fidelity measure (called the ACT-FM [54], with a sub-set of sessions evaluated by researchers. |  |
| **12. How well – actual**<br>Describe the extent to which the intervention was delivered as planned and outline the factors which had an impact on actual delivery | The intervention was successfully completed across three groups with excellent attendance and good adherence to protocol (fidelity). The staff training and support appeared to lead to good fidelity (detailed paper in progress). |  |
| **13. Voice**<br>Whose voice does this description of TIDieR convey?<br>Who was involved in the preparation of TIDieR, how they were involved in the intervention/their perspective (e.g. researcher, service deliverer, patient, etc.)? | This version of TIDieR represents the researcher's voice, with information gained through PPI included in the modifications column. |  |
| **14. Stage of implementation**<br>(i) What stage of implementation does the TIDieR checklist cover?<br>(ii) Is this a revision of an earlier TIDieR checklist | (i) Intervention development<br>(ii) No |  |

## References

1. The Stroke Association. Lived Experience of Stroke 2019; Available from: https://www.stroke.org.uk/lived-experience-of-stroke-report.

2. Lincoln, N.B., Psychological management of stroke. 2012, Chichester, UK: Wiley-Blackwell.

3. McKevitt, C., et al., Qualitative studies of stroke: a systematic review. Stroke, 2004. 35(6): p. 1499–1505.

4. Pollock, A., et al., Top ten research priorities relating to life after stroke. Lancet Neurology, 2012. 11(3): p. 209.

5. Stroke Association. Shaping stroke research to rebuild lives: The Stroke Priority Setting Partnership results for investment. 2021; Available from: https://www.stroke.org.uk/sites/default/files/research/stroke_priority_setting_partnership_full_report.pdf.

6. Sentinel Stroke National Audit Programme. Post-acute Organisational Audit National Report 2021; Available from: https://www.strokeaudit.org/Documents/National/PostAcuteOrg/2021/2021-PAOrgPublicReport.aspx.

7. NHS England, Guide to the provision of Psychological Support following Stroke. 2018.

8. Hayes, S.C., Acceptance and commitment therapy, relational frame theory, and the third wave of behavioral and cognitive therapies–republished article. Behavior therapy, 2016. 47(6): p. 869–885.

9. Graham, C.D., et al., An acceptance and commitment therapy (ACT)–based intervention for an adult experiencing post-stroke anxiety and medically unexplained symptoms. Clinical Case Studies, 2015. 14(2): p. 83–97.

10. Sathananthan, N., et al., A single-case experimental evaluation of a new group-based intervention to enhance adjustment to life with acquired brain injury: VaLiANT (valued living after neurological trauma). Neuropsychological Rehabilitation, 2022. 32(8): p. 2170–2202.

11. Rauwenhoff, J.C., et al., Acceptance and commitment therapy for individuals with depressive and anxiety symptoms following acquired brain injury: A non-concurrent multiple baseline design across four cases. Neuropsychological rehabilitation, 2022: p. 1–31.

12. Kangas, M. and S. McDonald, Is it time to act? The potential of acceptance and commitment therapy for psychological problems following acquired brain injury. Neuropsychological rehabilitation, 2011. 21(2): p. 250–276.

13. McLeod, H., Acceptance and commitment therapy’s value as a neuropsychotherapy. The Neuropsychologist, 2015. 1: p. 14–15.

14. Whiting, D.L., et al., Cognitive and psychological flexibility after a traumatic brain injury and the implications for treatment in acceptance-based therapies: A conceptual review. Neuropsychological Rehabilitation, 2017. 27(2): p. 263–299.

15. Niu, Y., et al., The efficacy of group acceptance and commitment therapy for preventing post-stroke depression: a randomized controlled trial. Journal of Stroke and Cerebrovascular Diseases, 2022. 31(2): p. 106225.

16. Whiting, D., et al., Can acceptance and commitment therapy facilitate psychological adjustment after a severe traumatic brain injury? A pilot randomized controlled trial. Neuropsychological rehabilitation, 2019.

17. Hill, G., et al., Living well with neurological conditions: Evaluation of an ACT-informed group intervention for psychological adjustment in outpatients with neurological problems. The Neuropsychologist, 2017. 3: p. 58–63.

18. Majumdar, S. and R. Morris, Brief group-based acceptance and commitment therapy for stroke survivors. British Journal of Clinical Psychology, 2019. 58(1): p. 70–90.

19. Burlingame, G.M., K. MacKenzie, and B. Strauss, Small group treatment: Evidence for effectiveness and mechanisms of change. Handbook of psychotherapy and behavior change. Vol. 5. 2004. 647–696.

20. Tucker, M. and T.P. Oei, Is group more cost effective than individual cognitive behaviour therapy? The evidence is not solid yet. Behavioural Cognitive Psychotherapy, 2007. 35(1): p. 77–91.

21. Ford, G.A., et al. Restoration and recovery of stroke services during the COVID-19 pandemic 2020; Available from: https://www.basp.org/supporting-stroke-services-during-the-covid-19-pandemic/restoration-and-recovery/.

22. NHS: Getting It Right First Time (GIRFT). Clinical guide for the management of virtual working in secondary care during the coronavirus pandemic. 2020; Available from: https://www.england.nhs.uk/coronavirus/wp-content/uploads/sites/52/2020/03/C0044-Specialty-Guide-Virtual-Working-and-Coronavirus-27-March-20.pdf.

23. Hill, G. and J. Price. Living Well with Neurological Conditions: An eight-week series of group workshops informed by Acceptance and Commitment Therapy (ACT). Protocol. 2017 [cited 2017; Available from: goo.gl/MBW12C.

24. Skivington, K., et al., A new framework for developing and evaluating complex interventions: update of Medical Research Council guidance. BMJ, 2021. 374.

25. Price, A., et al., Patient and public involvement in research: a journey to co-production. Patient Education and Counseling, 2022. 105(4): p. 1041–1047.

26. Nasreddine, Z.S., et al., Montreal cognitive assessment. The American Journal of Geriatric Psychiatry, 2003.

27. Enderby, P.M., et al., The Frenchay Aphasia Screening Test: a short, simple test for aphasia appropriate for non-specialists. International rehabilitation medicine, 1986. 8(4): p. 166–170.

28. Wilson, S.M., et al., A quick aphasia battery for efficient, reliable, and multidimensional assessment of language function. PloS one, 2018. 13(2): p. e0192773.

29. Enderby, P., A. John, and B. Petheram, Therapy outcome measures for rehabilitation professionals: speech and language therapy, physiotherapy, occupational therapy. 2013: John Wiley & Sons.

30. Zigmond, A.S. and R.P. Snaith, The hospital anxiety and depression scale. Acta psychiatrica scandinavica, 1983. 67(6): p. 361–370.

31. Benson, T., et al., Personal Wellbeing Score (PWS)—a short version of ONS4: development and validation in social prescribing. BMJ open quality, 2019. 8(2): p. e000394.

32. Collin, C., et al., The Barthel ADL Index: a reliability study. International disability studies, 1988. 10(2): p. 61–63.

33. Herdman, M., et al., Development and preliminary testing of the new five-level version of EQ-5D (EQ-5D-5L). Quality of life research, 2011. 20: p. 1727–1736.

34. Whiting, D.L., et al., Validating measures of psychological flexibility in a population with acquired brain injury. Psychological assessment, 2015. 27(2): p. 415.

35. Smout, M., et al., Development of the valuing questionnaire (VQ). Journal of Contextual Behavioral Science, 2014. 3(3): p. 164–172.

36. Suresh, K. and S. Chandrashekara, Sample size estimation and power analysis for clinical research studies. Journal of human reproductive sciences, 2012. 5(1): p. 7.

37. Frampton, G.K., et al., Digital tools for the recruitment and retention of participants in randomised controlled trials: a systematic map. Trials, 2020. 21(1): p. 1–23.

38. Clancy, B., et al., Access to and use of internet and social media by low-morbidity stroke survivors participating in a national web-based secondary stroke prevention trial: cross-sectional survey. Journal of Medical Internet Research, 2022. 24(5): p. e33291.

39. Clark, N., M. Lawrence, and B. Davis, Using social media to recruit stroke survivors to the HEADS: UP Online randomised control trial, a stroke-specific psychological self-management intervention: reflections and lessons learned. International Journal of Stroke, 2023. 18(1 supplement, 17th UK Stroke Forum Abstracts): p. 120–121.

40. Schwamm, L.H., et al., Race/ethnicity, quality of care, and outcomes in ischemic stroke. Circulation, 2010. 121(13): p. 1492–1501.

41. NHS England, The NHS Long Term Plan. 2019: www.longtermplan.nhs.uk.

42. Ghomi, M., et al., Development and validation of the Readiness for Therapy Questionnaire (RTQ). Behavioural and Cognitive Psychotherapy, 2021. 49(4): p. 413–425.

43. Ogrodniczuk, J.S., A.S. Joyce, and W.E. Piper, Development of the readiness for psychotherapy index. The Journal of nervous and mental disease, 2009. 197(6): p. 427–433.

44. Asensio, D. and J.A. Duñabeitia, The necessary, albeit belated, transition to computerized cognitive assessment. Frontiers in Psychology, 2023. 14: p. 1160554.

45. Zhang, Q., et al., Can we trust computers to assess the cognition of stroke patients? A systematic review. Frontiers in Neurology, 2023. 14: p. 1180664.

46. Gallacher, K.I., et al., Multimorbidity in stroke. Stroke, 2019. 50(7): p. 1919–1926.

47. Foote, H., et al., A scoping review to identify process and outcome measures used in acceptance and commitment therapy research, with adults with acquired neurological conditions. Clinical Rehabilitation, 2023. 37(6): p. 808–835.

48. Lemay, K.R., et al., Establishing the minimal clinically important difference for the hospital anxiety and depression scale in patients with cardiovascular disease. Journal of cardiopulmonary rehabilitation and prevention, 2019. 39(6): p. E6–E11.

49. Craig, P., et al., Developing and evaluating complex interventions: the new Medical Research Council guidance. BMJ, 2008. 337: p. a1655.

50. Hoffmann, T.C., et al., Better reporting of interventions: template for intervention description and replication (TIDieR) checklist and guide. BMJ 2014. 348.

51. Cotterill, S., et al., Getting messier with TIDieR: embracing context and complexity in intervention reporting. BMC medical research methodology, 2018. 18(1): p. 12.

52. Graham, C.D., et al., A systematic review of the use of Acceptance and Commitment Therapy (ACT) in chronic disease and long-term conditions. Clinical psychology review, 2016. 46: p. 46–58.

53. Marshall, R.S., J. Laures-Gore, and K. Love, Brief mindfulness meditation group training in aphasia: Exploring attention, language and psychophysiological outcomes. International journal of language communication disorders, 2018. 53(1): p. 40–54.

54. O’Neill, L., et al., The development of the Acceptance and Commitment Therapy Fidelity Measure (ACT-FM): A delphi study and field test. Journal of Contextual Behavioral Science, 2019. 14: p. 111–118.

